# Protective Dietary Antioxidants Intake Attenuate Cardiovascular-Kidney-Metabolic Syndrome Progression

**DOI:** 10.1101/2025.08.01.25332845

**Authors:** Xiangyun Dang, Danfeng Ren, Fangang Zeng, Guiqing Xu, Zhenpeng Zhou, Yafeng Zhao, Nan Zhang, Zhongxin Jin, Ting Li, Liang Ma, Jinfeng Liu, Xiaoyong Yu

**Author notes:** These authors have contributed equally to this work and share first authorship (Xiangyun Dang, Danfeng Ren). Co-corresponding: Xiaoyong Yu, M.D., Department of Nephrology, Shaanxi Provincial Hospital of Chinese Medicine, No. 4, Xihuamen Street, Lianhu District, Xi’an, Shaanxi, 710003, China, Jinfeng Liu, Ph.D., Department of Infectious Diseases, The First Affiliated Hospital of Xi’an Jiaotong University, No. 277, West Yanta Road, Yanta District, Xi’an, Shaanxi, 710061, China.

## Abstract

**Background:** Cardiovascular-kidney-metabolic (CKM) syndrome is characterized by complex pathophysiological interactions among cardiovascular diseases, and chronic kidney disease. Although there is evidence linking dietary antioxidants to the reduction of oxidative stress, comprehensive studies investigating relationships between CKM syndrome and antioxidants remain limited.

**Methods:** This cross-sectional study analyzed data from 5,349 participants (NHANES 2007–2010, 2017–2018) to evaluate the associations between 43 dietary antioxidants and CKM syndrome stages. Participants were categorised into subgroups: CKM syndrome vs. non-CKM syndrome and CKM syndrome stage groups (0-4). Sequential ordinal logistic regression was employed while adjusting for demographics, socioeconomic status, and lifestyle variables. Five machine learning models—XGBoost, Balanced Random Forest (BRF), Support Vector Machine (SVM), Glmnet, and Artificial Neural Network (ANN)—were trained after removing multicollinearity features to identify predictors for CKM syndrome. Model performance was assessed using AUC-ROC, sensitivity, specificity metrics, and SHAP analysis for interpretability enhancement.

**Results:** The CKM syndrome group (90.8% of participants) exhibited lower intakes of daidzein (0.663 vs. 1.494 mg/day, *p =* 0.004) and genistein (0.976 vs. 2.149 mg/day, *p =* 0.007) compared to non-CKM syndrome. There were significant statistical differences in age, sex, race-ethnicity, education level, family income-to-poverty ratio, weekly physical activity status, and smoking status among the 4 stages of CKM syndrome (all *p*<0.05). Ordinal regression revealed significant inverse associations between CKM syndrome progression and antioxidants: vitamin C (OR: 0.999, *p =* 0.041), magnesium (OR: 0.999, *p*<0.001), daidzein (OR: 0.965, *p =* 0.047), genistein (OR: 0.977, *p =* 0.037), delphinidin (OR: 0.983, *p =* 0.024), apigenin (OR: 0.952, *p*<0.001), luteolin (OR: 0.930, *p =* 0.031), kaempferol(OR:0.986, *p =* 0.026), total flavones(OR: 0.958, *p =* 0.012), and total flavonols(OR: 0.996, *p =* 0.049). ML models identified XGBoost as optimal (AUC-ROC: 0.922, specificity: 0.852), with age as the strongest risk predictor (SHAP: 0.535). Protective antioxidants included selenium (−0.1838), luteolin (−0.2019), total flavonoids (−0.1345), vitamin E (−0.1511), magnesium (−0.1849), vitamin A (−0.1987), myricetin (−0.1483), vitamin C (−0.1862), kaempferol (−0.1546), zinc (−0.1615), subtotal catechins (−0.1172), and total flavan-3-ols (−0.2073).

**Conclusion:** Dietary antioxidants, particularly flavonoids and vitamins, exhibit protective associations against the progression of CKM syndrome. These findings support the implementation of targeted dietary interventions and advocate for early screening in high-risk populations.

## 1. Introduction

Cardiovascular-kidney-metabolic (CKM) syndrome is a novel systemic ailment characterized by intricate pathophysiological interactions among subclinical or clinical cardiovascular disease (CVD), chronic kidney disease (CKD), and metabolic disorders[1–4]. The American Heart Association (AHA) has classified CKM syndrome into stages 0 to 4[5].In the United States, over one in four adults is affected by at least one condition within this triad[6], with only 10.6% of adults aged ≥20 years being free from CKM syndrome, predominantly affecting those in stages 1 and 2[7]. Globally, approximately 25% to 30% of individuals are impacted by concurrent comorbidities associated with CKM syndrome[6]. The burden imposed by the CKM syndrome on both patients and healthcare systems is substantial.

Previous studies have demonstrated that the intake levels of vitamins C and E, polyphenols, and carotenoids can effectively alleviate oxidative stress, a well-established pathological pathway in CKM syndrome[8–10]. There existed an inverse relationship between vitamin E intake and the risk of CVD. The total antioxidant capacity of the diet was found to be inversely associated with cardiovascular events and cardiometabolic risk factors in individuals with a history of CVD[11, 12]. It has been suggested that increased dietary intake of flavonoids is associated with enhanced cardiovascular outcomes, protection against diabetes and obesity, as well as a deceleration in the progression of chronic kidney disease (CKD)[12–15]. However, significant gaps remain in our understanding of the relationship between CKM syndrome and dietary antioxidant intake. Existing research primarily focused on the association between antioxidants and a single CKM syndrome condition, overlooking the effects that may arise from the antioxidants across this spectrum. Moreover, there is a notable deficiency of comprehensive studies that encompass the full range of stages within CKM syndrome alongside various types of dietary antioxidants. Additionally, relatively few investigations have explored the substantial relationship between dietary antioxidants and CKM syndrome while considering the modifying effects of socioeconomic and lifestyle factors. This study aimed to identify nonlinear thresholds and synergistic interactions among antioxidants by employing machine learning (ML) models, thereby elucidating the relationship between dietary antioxidant features and CKM syndrome. Our objective was to provide more robust evidence regarding the potential role of dietary antioxidants in managing CKM syndrome and to delineate specific dietary recommendations that could enhance clinical outcomes.

## Materials and Methods

### 2.1 Study Design

Data for this cross-sectional study were obtained from three cycles (2007–2008, 2009-2010 and 2017-2018) of the National Health and Nutrition Examination Survey (NHANES, https://www.cdc.gov/nchs/nhanes). This survey encompassed demographic, socioeconomic, dietary, and health-related information from the US population. Participants were interviewed at home and underwent standardized physical examinations and biochemical sample collections in mobile examination centres (MEC) every two years. The NHANES protocols received approval from the Research Ethics Review Board of the National Center for Health Statistics, with written informed consent obtained from all participants. This retrospective study followed the Strengthening the Reporting of Observational Studies in Epidemiology (STROBE) guidelines. Among 22,483 eligible participants, 215 pregnant individuals and 6,051 individuals with ages < 20 and > 79 years were initially excluded. In addition, 10,868 participants who lacked flavonoid intake values or information regarding CKD diagnosis were excluded. Consequently, a total of 5,349 participants with complete information were enrolled in this study **(Figure 1)**.

**Figure 1.**
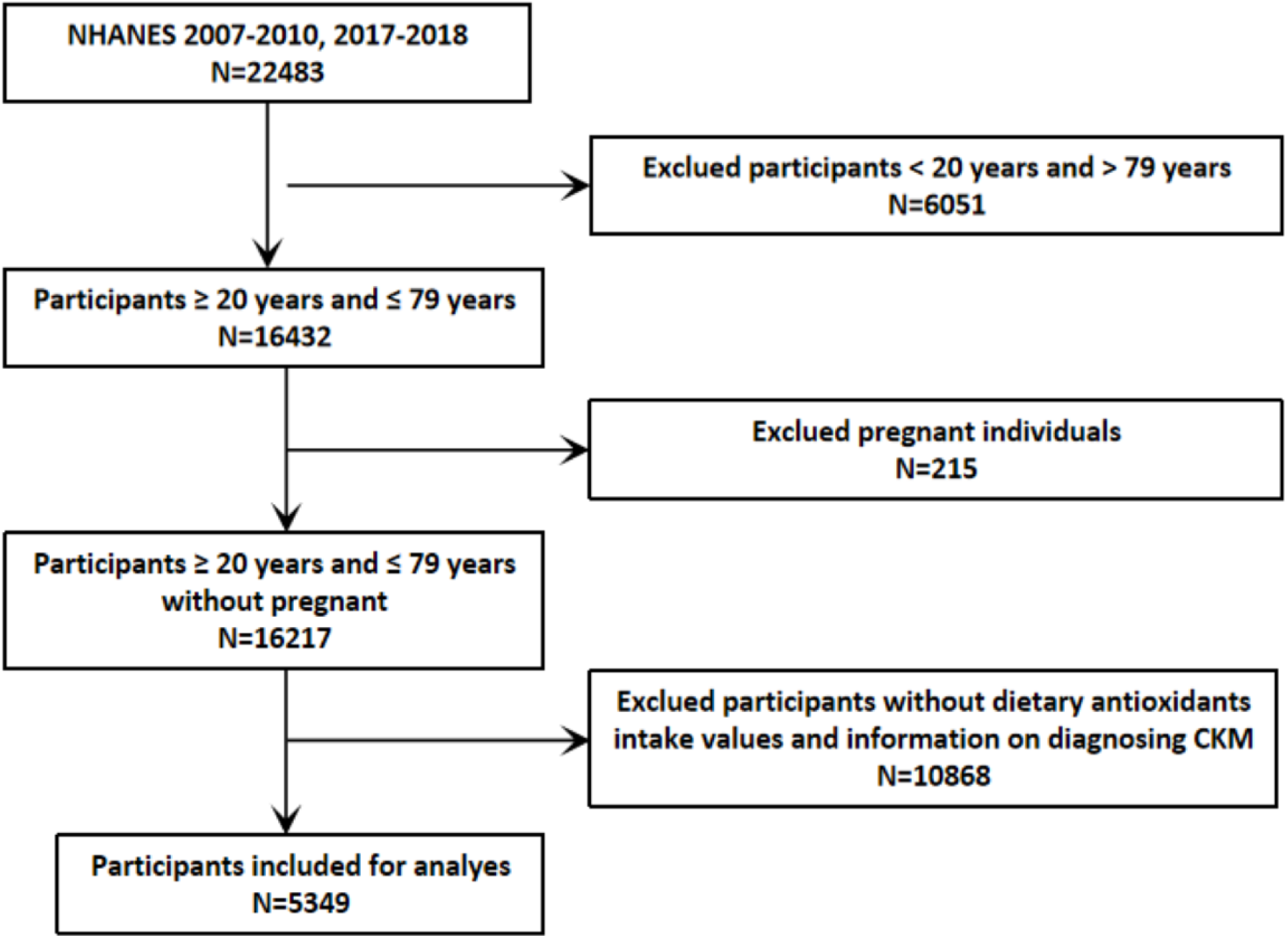
Flowchart for participant screening.

### 2.2 Baseline Data Collection

The variables of age, sex, race-ethnicity, education level, the ratio of family income to poverty, weekly moderate to vigorous physical activity status, and current smoking habits were acquired from self-reported questionnaires. Race-ethnicity variables were categorized into Mexican American, other Hispanic, non-Hispanic white, non-Hispanic black, non-Hispanic Asian and other races (including multi-racial). Education levels were classified as less than high school diploma, high school graduate, some college or associate degree holders, and those with a college degree or higher. The ratio of family income to poverty was simplified into ≤3 and >3. Both weekly moderate to vigorous activity status and current smoking status were classified as either “yes” or “no.”

### 2.3 Antioxidants Intake Data

The dietary intake data in NHANES were collected through two 24-hour dietary recall interviews conducted at the Mobile Examination Center (MEC). The time interval between these two interviews ranged from 3 to 10 days. Data on the intake of 43 dietary antioxidants were obtained, which includes 6 vitamins and minerals sourced as well as 37 flavonoid values for all foods and beverages listed in the USDA Food and Nutrient Database for Dietary Studies (FNDDS) and NHNAES. Subsequently, the average daily intake of dietary antioxidants was calculated based on the collected dietary intake data.

### 2.4. Diagnostic Criteria of CKM Syndrome

The detailed definition of CKM syndrome was presented in **Table Supplement 1.** Clinical CVD included ischemic heart disease and cerebrovascular diseases. Subclinical CVD was characterized by a 10-year CVD risk of ≥20% or classification as high-risk for CKD. The 10-year predicted CVD risk was calculated utilizing the simplified PREVENT equations algorithm[7, 16, 17], which incorporates variables including age, sex, tobacco use, blood pressure, cholesterol levels, diabetes status, kidney function, antihypertensive medication usage, and statin therapy within its model[16]. In this study, CKD was defined according to moderate– or high-risk classifications based on the Kidney Disease: Improving Global Outcomes guidelines[18, 19]. Metabolic disorders include conditions such as overweight/obesity, abdominal obesity, prediabetes, diabetes, hypertension, dyslipidemia, and metabolic syndrome.

Participants were categorised into CKM syndrome vs. non-CKM syndrome subgroups. In addition, individuals were classified into five stages on the varying clinical severities of different manifestations of CKM syndrome[7, 16]: Stage 0, absence of CKM syndrome risk factors; Stage 1: with overweight/obesity, abdominal obesity, or dysfunctional adipose tissue, without the presence of metabolic risk factors associated with CKD; Stage 2: exhibiting metabolic risk factors or CKD; Stage 3: presenting subclinical CVD alongside metabolic risk factors or CKD; stage 4: involving clinical CVD combined metabolic disorders or CKD[5]. The detailed criteria for each stage were shown in **Table Supplement 2.** Additionally, we divided participants into two subgroups for further machine learning analysis: the CKM syndrome group (Stages 1-4) and the non-CKM syndrome group (Stage 0).

### 2.5 Statistical Analysis

#### 2.5.1 Descriptive Statistics

Continuous variables were presented as mean ± standard deviation (SD), whereas categorical variables are expressed as numbers (percentages). Inter-group comparisons were performed using weighted Student’s t-tests or Kruskal-Wallis test for continuous variables, and weighted chi-square tests for categorical variables.

#### 2.5.2 Multivariable Ordinal Logistic Regression

Three ordinal logistic regression models were constructed to explore the independent association between dietary antioxidant features and CKM syndrome: Model 1 was unadjusted, while Model 2 was adjusted for age, gender, and race.

Model 3 included further adjustments for family income-to-poverty ratio, education level, moderate-to-vigorous physical activity, and smoking status. Odds Ratios (OR) along with 95% confidence intervals (CIs) were documented.

#### 2.5.3 Machine Learning Models

To address multicollinearity among 43 dietary antioxidant features, pairwise Spearman correlations were calculated. Features with correlation coefficients >0.9 were excluded. Furthermore, the labels and results presented in this study pertain to binary or multi-class classification data. Therefore, it seemed more suitable to employ a classification algorithm in a supervised learning framework[20]. Subsequently, five ML algorithms were implemented using the mlr3 platform: Support Vector Machine (SVM), Balanced Random Forest (BRF), eXtreme Gradient Boosting (XGBoost), Generalized Linear Models Network (Glmnet), and Artificial Neural Network (ANN). The SVM model identifies an optimal hyperplane in high-dimensional spaces, making it well-suited for non-linear classification and regression tasks[21]. BRF builds upon the random forest algorithm and is specifically designed to address class imbalance issues. XGBoost is an optimized gradient boosting framework that uses second-derivative information to speed up convergence, allowing for efficient processing of large datasets[22]. Glmnet is employed for fitting generalized linear models while conducting variable selection and implementing regularization techniques. ANN, inspired by biological neural networks, models complex non-linear relationships through its interconnected structure[23]. The class imbalance between the CKM syndrome and non-CKM syndrome groups was addressed using the Synthetic Minority Over-sampling Technique (SMOTE), which generates synthetic minority samples through linear interpolation of the k=7 nearest neighbors. All features were standardized using z-score transformation (mean=0, SD=1) to minimize scale-induced bias during model training.

#### 2.5.4 SHAP Analysis

We performed a SHAP (SHapley Additive exPlanations) analysis to evaluate feature importance in the optimally performing machine learning model. SHAP is an advanced interpretability framework grounded in cooperative game theory, which assesses the marginal contributions of features by examining all possible permutations. This methodology provides both global rankings of feature importance and instance-level explanatory visualizations.

#### 2.5.5 Statistical Software

Data analysis and figure plot were conducted using the R software(version 4.4.3). The following R packages were employed for statistical analysis: nhanesA, dplyr, survey, MASS, mlr3, mlr3pipelines, mlr3extralearner, mlr3tuning, mlr3filters, shapviz, corrplot, ggplot2, and pheatmap. All statistical tests performed were two-sided, with a p-value < 0.05 considered statistically significant.

## 2. Results

### 3.1. Characteristics of Participants and Dietary Antioxidant Features

Among a total of 5,349 individuals included, there were 4,858 (90.82%) and 715 (9.18%) individuals in CKM syndrome and non-CKM syndrome groups, respectively. The CKM syndrome group exhibited a higher proportion of older adults (≥ 65 years: 16.39% vs. 4.93%, *p* < 0.0001) and males (53.82% vs. 32.75%, *p* < 0.0001). A significantly lower percentage of CKM syndrome participants engaged in moderate-to-vigorous physical activity every week compared to their non-CKM syndrome counterparts (25.77% vs. 39.95%, *p* < 0.0001). The CKM syndrome group also demonstrated a lower intake of daidzein (0.663 vs. 1.494 mg/day, *p* = 0.004) and genistein (0.976 vs. 2.149 mg/day, *p* = 0.007). Table 1 summarizes the characteristics of all features between the CKM syndrome and non-CKM syndrome groups.

**Table 1.** Characteristics of participants in the CKM syndrome and non-CKM syndrome groups from NHANES 2007-2010 and 2017-2018. Continuous variables are presented as mean±SD, while categorical variables are shown as number (%).

| Variables | CKM syndrome | Non-CKM syndrome | *p*-value |
| --- | --- | --- | --- |
| Vitamin A(mg/day) | 635.982±13.111 | 645.645±21.678 | 0.695 |
| Vitamin C(mg/day) | 80.738±1.654 | 78.263±2.845 | 0.426 |
| Vitamin E(mg/day) | 8.329±0.135 | 8.982±0.322 | 0.060 |
| Mg(mg/day) | 305.222±4.518 | 302.742±6.945 | 0.764 |
| Zn(mg/day) | 11.978±0.192 | 11.622±0.336 | 0.412 |
| Se(mg/day) | 116.867±1.432 | 112.487±3.379 | 0.287 |
| Daidzein(mg/day) | 0.663±0.084 | 1.494±0.255 | 0.004 |
| Genistein(mg/day) | 0.976±0.146 | 2.149±0.38 | 0.007 |
| Glycitein(mg/day) | 0.145±0.023 | 0.302±0.055 | 0.013 |
| Cyanidin(mg/day) | 2.493±0.195 | 2.260±0.249 | 0.466 |
| Petunidin(mg/day) | 1.101±0.107 | 1.387±0.276 | 0.345 |
| Delphinidin(mg/day) | 1.540±0.150 | 2.013±0.373 | 0.241 |
| Malvidin(mg/day) | 5.193±0.513 | 6.013±0.913 | 0.439 |
| Pelargonidin(mg/day) | 1.485±0.136 | 1.544±0.221 | 0.789 |
| Peonidin(mg/day) | 1.717±0.175 | 2.812±0.590 | 0.107 |
| Catechin(mg/day) | 8.035±0.241 | 8.129±0.410 | 0.834 |
| Epigallocatechin(mg/day) | 17.499±0.910 | 15.579±1.407 | 0.252 |
| Epicatechin(mg/day) | 10.337±0.286 | 10.115±0.495 | 0.702 |
| Epicatechin 3-gallate(mg/day) | 11.221±0.600 | 10.221±0.952 | 0.364 |
| Epigallocatechin 3-gallate(mg/day) | 29.862±2.025 | 26.927±2.533 | 0.390 |
| Theaflavin(mg/day) | 1.665±0.109 | 1.556±0.168 | 0.526 |
| Thearubigins(mg/day) | 94.764±5.875 | 87.860±8.941 | 0.475 |
| Eriodictyol(mg/day) | 0.172±0.016 | 0.148±0.020 | 0.366 |
| Hesperetin(mg/day) | 8.351±0.415 | 7.323±0.562 | 0.165 |
| Naringenin(mg/day) | 3.055±0.191 | 2.796±0.273 | 0.378 |
| Apigenin(mg/day) | 0.254±0.049 | 0.271±0.061 | 0.820 |
| Luteolin(mg/day) | 0.743±0.032 | 0.724±0.043 | 0.707 |
| Isorhamnetin(mg/day) | 0.885±0.036 | 0.920±0.080 | 0.616 |
| Kaempferol(mg/day) | 4.941±0.130 | 4.569±0.385 | 0.315 |
| Myricetin(mg/day) | 1.636±0.057 | 1.424±0.089 | 0.057 |
| Quercetin(mg/day) | 11.766±0.277 | 11.179±0.502 | 0.207 |
| Theaflavin 3-3 digallate(mg/day) | 1.834±0.121 | 1.715±0.186 | 0.533 |
| Theaflavin 3-q gallate(mg/day) | 1.558±0.103 | 1.460±0.159 | 0.545 |
| Theaflavin 3-gallate(mg/day) | 1.317±0.087 | 1.230±0.133 | 0.530 |
| Gallocatechin(mg/day) | 1.798±0.086 | 1.675±0.155 | 0.449 |
| Subtotal Catechins(mg/day) | 107.424±4.052 | 96.904±8.894 | 0.249 |
| Total Isoflavones(mg/day) | 3.968±0.365 | 6.364±1.156 | 0.074 |
| Total Anthocyanidins(mg/day) | 26.487±1.740 | 25.645±2.987 | 0.802 |
| Total Flavan-3-ols(mg/day) | 179.892±8.465 | 166.466±14.528 | 0.382 |
| Total Flavanones(mg/day) | 11.578±0.550 | 10.268±0.728 | 0.159 |
| Total Flavones(mg/day) | 0.996±0.069 | 0.995±0.088 | 0.992 |
| Total Flavonols(mg/day) | 19.228±0.422 | 18.093±0.911 | 0.186 |
| Total Sum of all 29 flavonoids(mg/day) | 183.195±8.974 | 179.501±13.296 | 0.796 |
| Age(n, %) |  |  | < 0.0001 |
| <65 | 3814(83.607) | 670(95.075) |  |
| ≥65 | 1044(16.393) | 45(4.925) |  |
| Gender(n, %) |  |  | < 0.0001 |
| Female | 2286(46.182) | 504(67.249) |  |
| Male | 2572(53.818) | 211(32.751) |  |
| Race(n, %) |  |  | 0.101 |
| Mexican American | 859(9.431) | 119(7.371) |  |
| Non-Hispanic Black | 547(6.068) | 68(4.716) |  |
| Non-Hispanic White | 2064(65.223) | 349(70.175) |  |
| Other Hispanic | 992(11.431) | 107(9.270) |  |
| Other Race - Including Multi-Racial | 396(7.848) | 72(8.467) |  |
| Education(n, %) |  |  | 0.003 |
| Less than high school | 1251(16.414) | 122(9.500) |  |
| High school graduate | 1137(24.925) | 165(24.935) |  |
| Some college or associates degree | 1422(30.156) | 217(30.024) |  |
| College or above | 1048(28.505) | 211(35.541) |  |
| Ratio of family income to poverty level(n, %) |  |  | 0.072 |
| ≤3 | 2901(48.996) | 396(44.390) |  |
| >3 | 1957(51.004) | 319(55.610) |  |
| Moderate to vigorous activity(n, %) |  |  | < 0.0001 |
| No | 3827(74.232) | 479(60.052) |  |
| Yes | 1031(25.768) | 236(39.948) |  |
| Current smoking(n, %) |  |  | 0.180 |
| No | 3884(81.411) | 550(77.891) |  |
| Yes | 974(18.589) | 165(22.109) |  |

The comparison of characteristics across CKM syndrome stages (0-4) was shown in Table 2. The proportion of participants aged ≥ 65 years increased sharply from 4.9% in stage 0 to 48.1% in stage 4 (*p* < 0.0001). Males were significantly more prevalent in the advanced stages (stage 4: 49.7% vs. stage 0: 32.8%, *p* < 0.0001). The percentage of individuals with less than a high school education increased from 9.5% (stage 0) to 29.4% (stage 4, *p* < 0.0001). Meanwhile, those with a family income-to-poverty ratio ≤3 increased from 44.4% in stage 0 to 59.4% in stage 4 (*p* = 0.003). Weekly moderate-to-vigorous activity rates markedly declined from 39.9% (stage 0) to 8.5% (stage 4, *p* < 0.0001), while the prevalence of current smoking peaked at stage 4 (26.2% vs. 22.1% at stage 0, *p* < 0.0001). The intake of isoflavones demonstrated a progressive decline with CKM syndrome advancement. daidzein intake significantly decreased from 1.494 ± 0.255 mg/day in stage 0 to 0.404 ± 0.118 mg/day in stage 4 (*p* < 0.0001), while genistein intake exhibited a corresponding reduction from 2.149 ± 0.380 mg/day to 0.519 ± 0.160 mg/day across the same stages (*p* < 0.0001). delphinidin intake also showed a significant decrease from stage 0 to stage 4 (2.013 ± 0.373 mg/day vs. 1.304 ± 0.351 mg/day, *p* = 0.013). Total isoflavones intake followed a similar declining trend from 6.364 ± 1.156 μg/day in stage 0 to 3.154 ± 0.684 mg/day in stage 3(*p* = 0.012). Although Vitamin E and Zinc intake displayed nonlinear trajectories across stage 0 to stage 4, significant differences were observed among stage subgroups(*p* < 0.001 and *p* = 0.040, respectively).

**Table 2.**
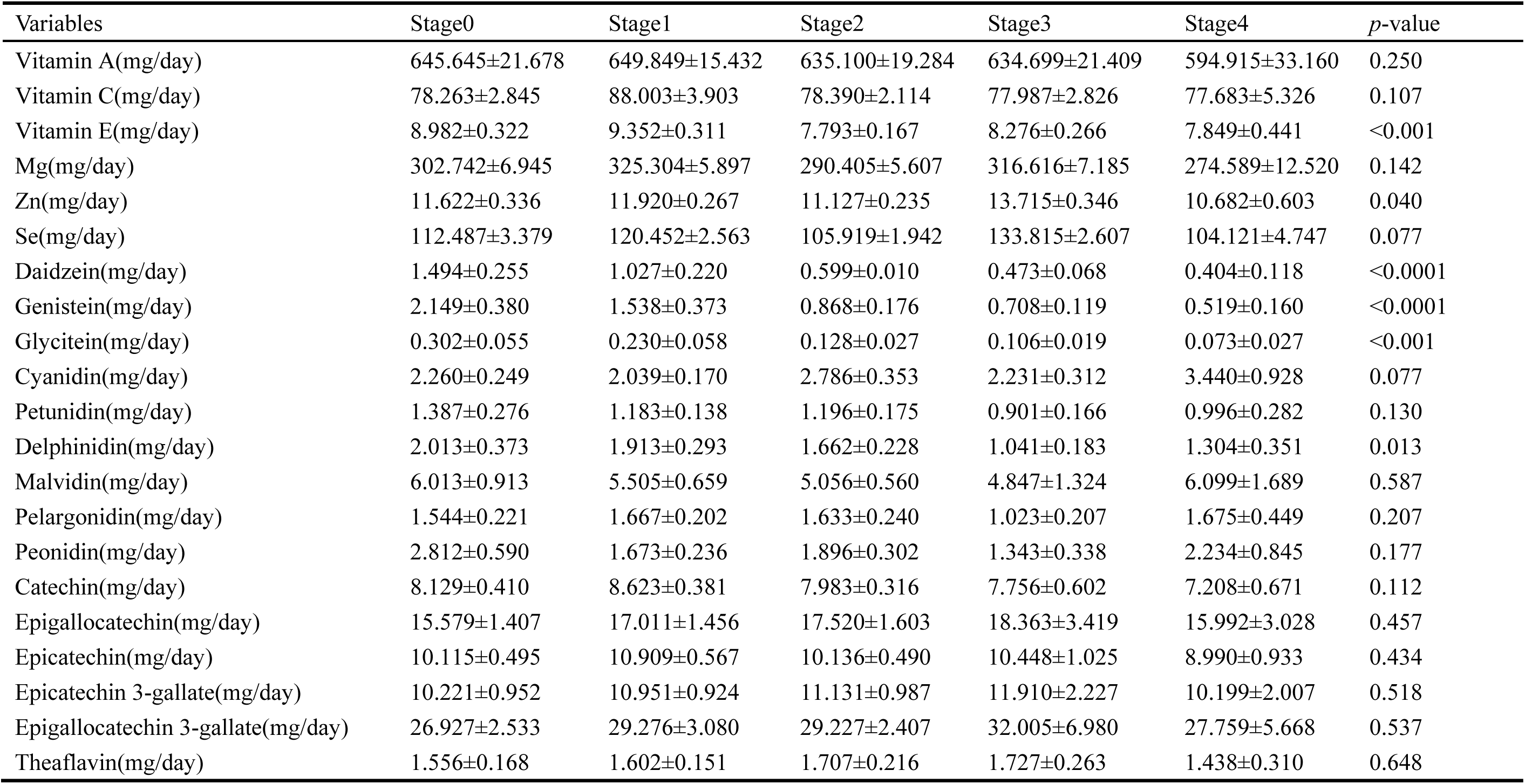
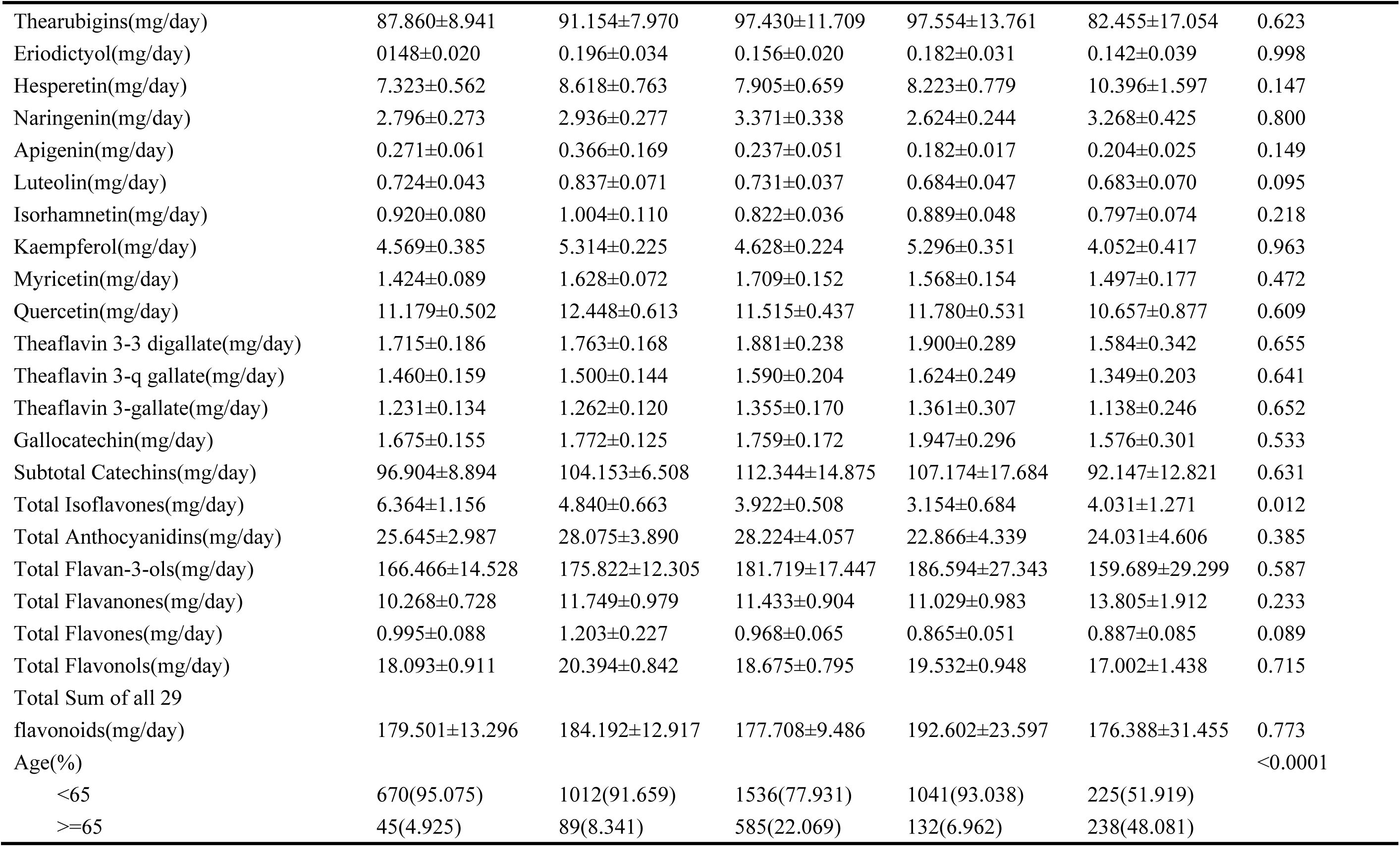
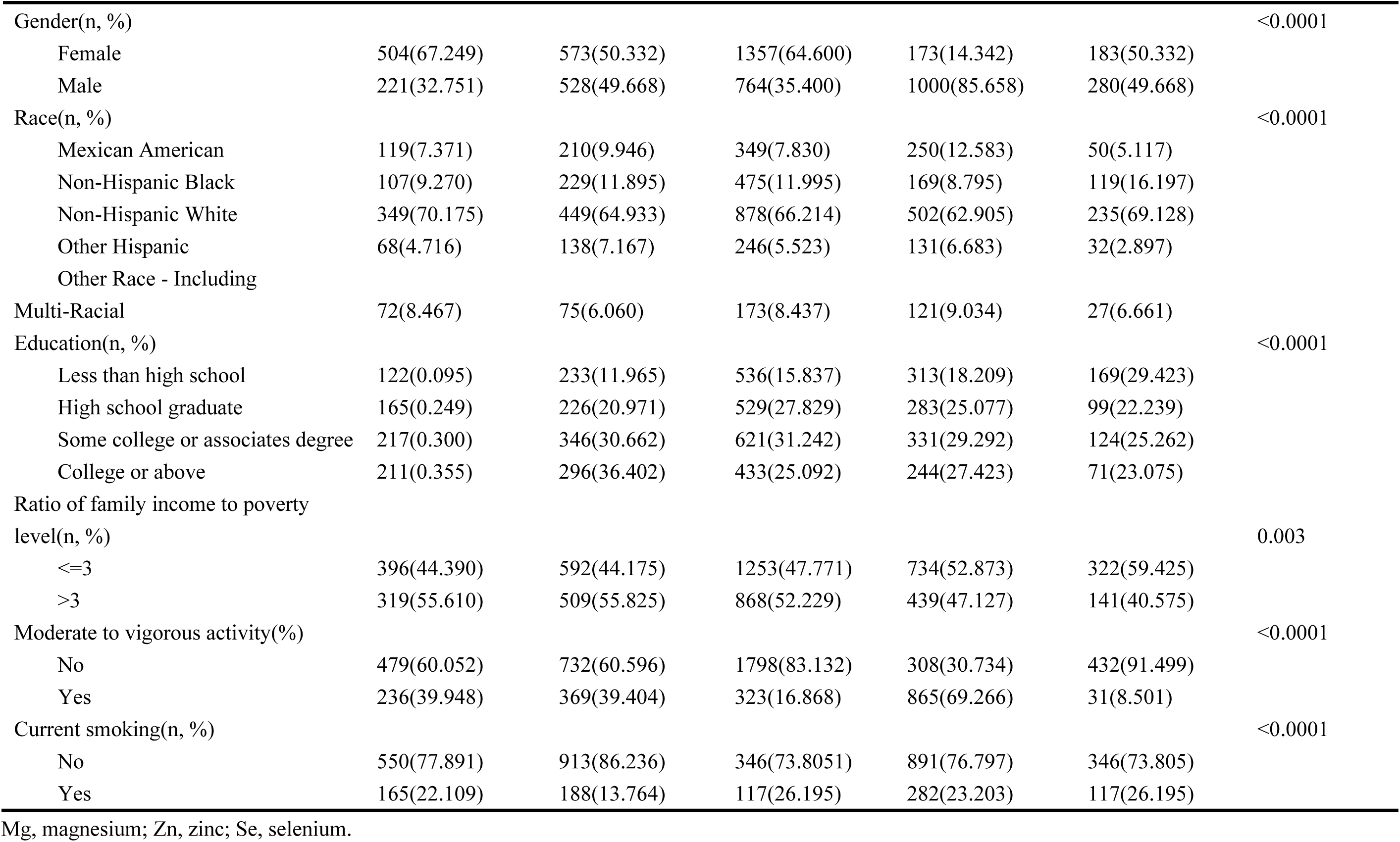
Characteristics of participants in the CKM syndrome stage (0-4) groups from NHANES 2007-2010 and 2017-2018. Continuous variables are presented as mean±SD, while categorical variables are shown as number (%).

### 3.3 Associations Between Dietary Antioxidant Features and CKM Syndrome

The associations between dietary antioxidant features and the progression of CKM syndrome (stages 0–4) were illustrated using ordinal logistic regression models, as presented in **Table 3**. The results indicated a significant inverse relationship between CKD stage and levels of various antioxidants: vitamin C (OR: 0.999, 95% CI: 0.998, 1.000, *p* = 0.041), magnesium (Mg) (OR: 0.999, 95% CI: 0.998, 0.999, *p* < 0.001), daidzein (OR: 0.965, 95% CI: 0.930-0.998, *p* = 0.047), genistein (OR: 0.977, 95% CI: 0.955, 0.999, *p* = 0.037), delphinidin (OR: 0.983, 95% CI: 0.962, 1.005, *p* = 0.024), apigenin (OR: 0.952, 95% CI: 0.926, 0.979, *p* < 0.001), luteolin (OR: 0.930, 95% CI: 0.842, 1.027, *p* = 0.031), kaempferol(OR: 0.986, 95% CI: 0.974, 0.998, *p* = 0.026), total flavones(OR: 0.958, 95% CI: 0.926, 0.991, *p* = 0.012), and total flavonols(OR: 0.996, 95% CI: 0.991,0.999, *p* = 0.049). These associations remained statistically significant even after adjusting for multiple covariates.

**Table 3.**
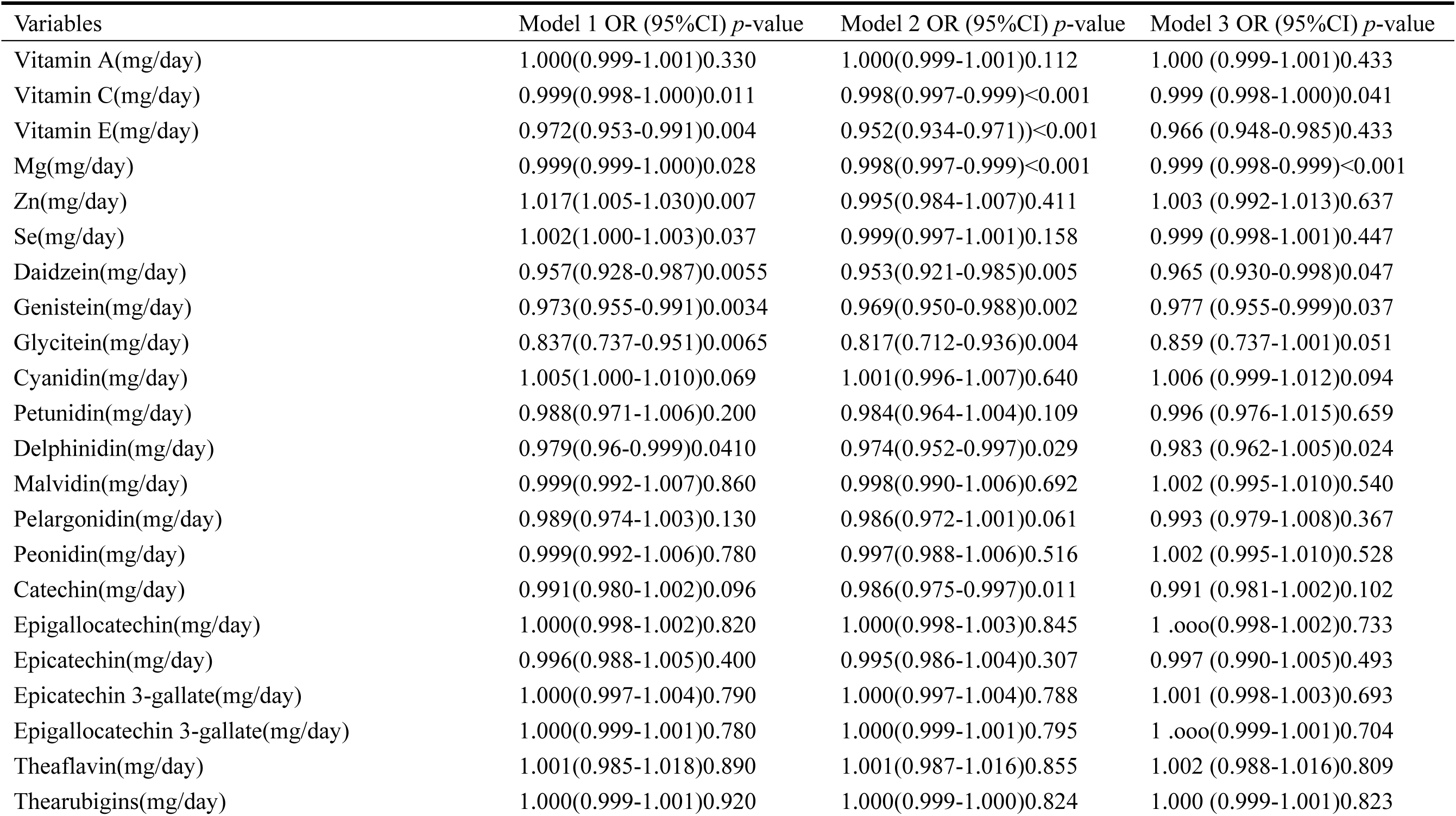
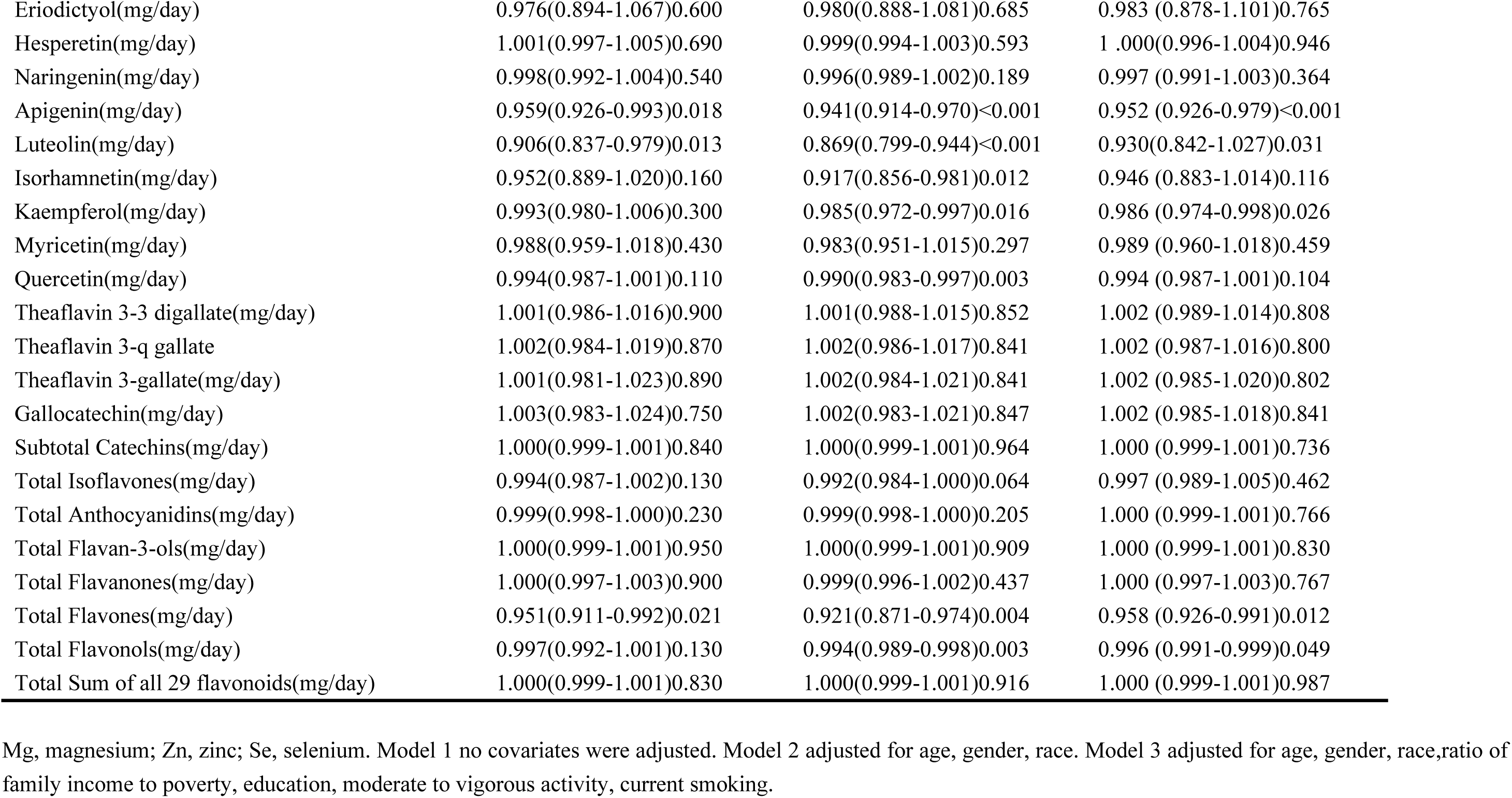
Associations between dietary antioxidants intake and CKM syndrome in three ordinal logistic regression models.

### 3.4 Machine Learning Models for Identifying Dietary Antioxidant Features with Stronger Associations to CKM Syndrome

Spearman correlation analysis showed significant positive correlations among various dietary antioxidant features (**Figure 2**). After excluding 14 features due to multicollinearity, 36 optimized features (30 dietary antioxidants and 7 demographic and lifestyle characteristics) were selected for ML model development.

**Figure 2.**
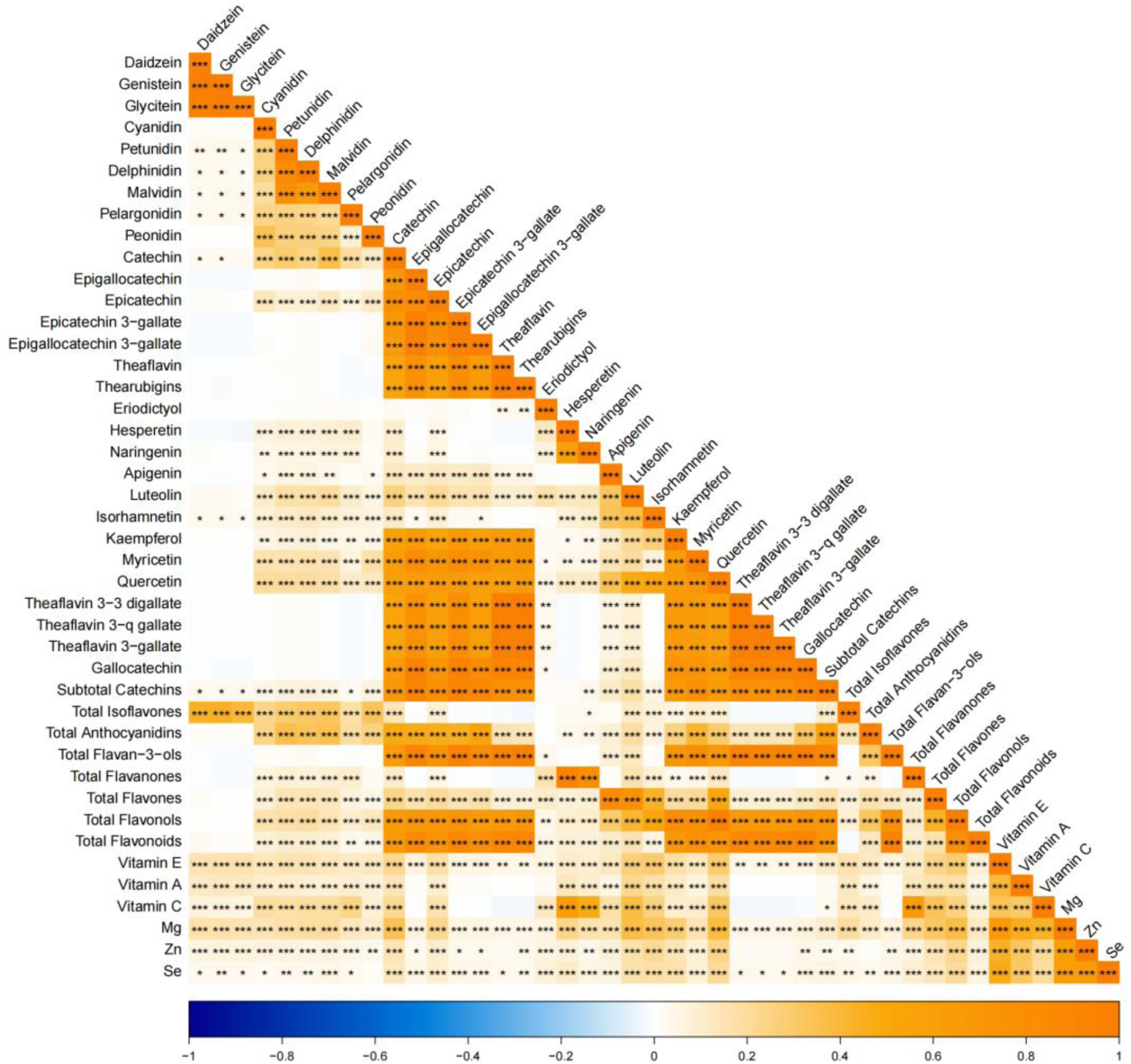
Correlations among dietary antioxidant features.

Five ML algorithms (SVM, BRF, XGBoost, Glmnet, and ANN) were implemented to identify key predictors of CKM syndrome outcomes. A comprehensive evaluation of performance metrics was conducted, including accuracy, F-beta score, AUC-ROC, sensitivity, specificity, and AUC-PR **(Table 4**; **Figure 3A and 3B).** Comparative analysis revealed statistically significant differences in performance across all models (*p* < 0.001). The XGBoost algorithm achieved the highest enhanced score (0.893) and AUC-ROC (0.922), along with robust specificity (0.852) and AUC-PR (0.960). BRF showed competitive performance with an F-beta score of 0.898 and an AUC-PR of 0.954 while maintaining high accuracy (0.851) and sensitivity (0.940). ANN exhibited moderate efficacy with an AUC-ROC of 0.813 and a specificity of 0.695. The comparative analysis indicated that Glmnet performed the worst across all metrics (accuracy: 0.729; AUC-ROC: 0.748), while SVM displayed notable sensitivity (0.929) but low specificity (0.523). Notably, both BRF and XGBoost demonstrated an optimal balance between sensitivity and specificity. Subsequently, we employed SHAP value analysis to interpret feature importance in the XGBoost model due to its excellent generalizability for clinical applications.

**Figure 3A.**
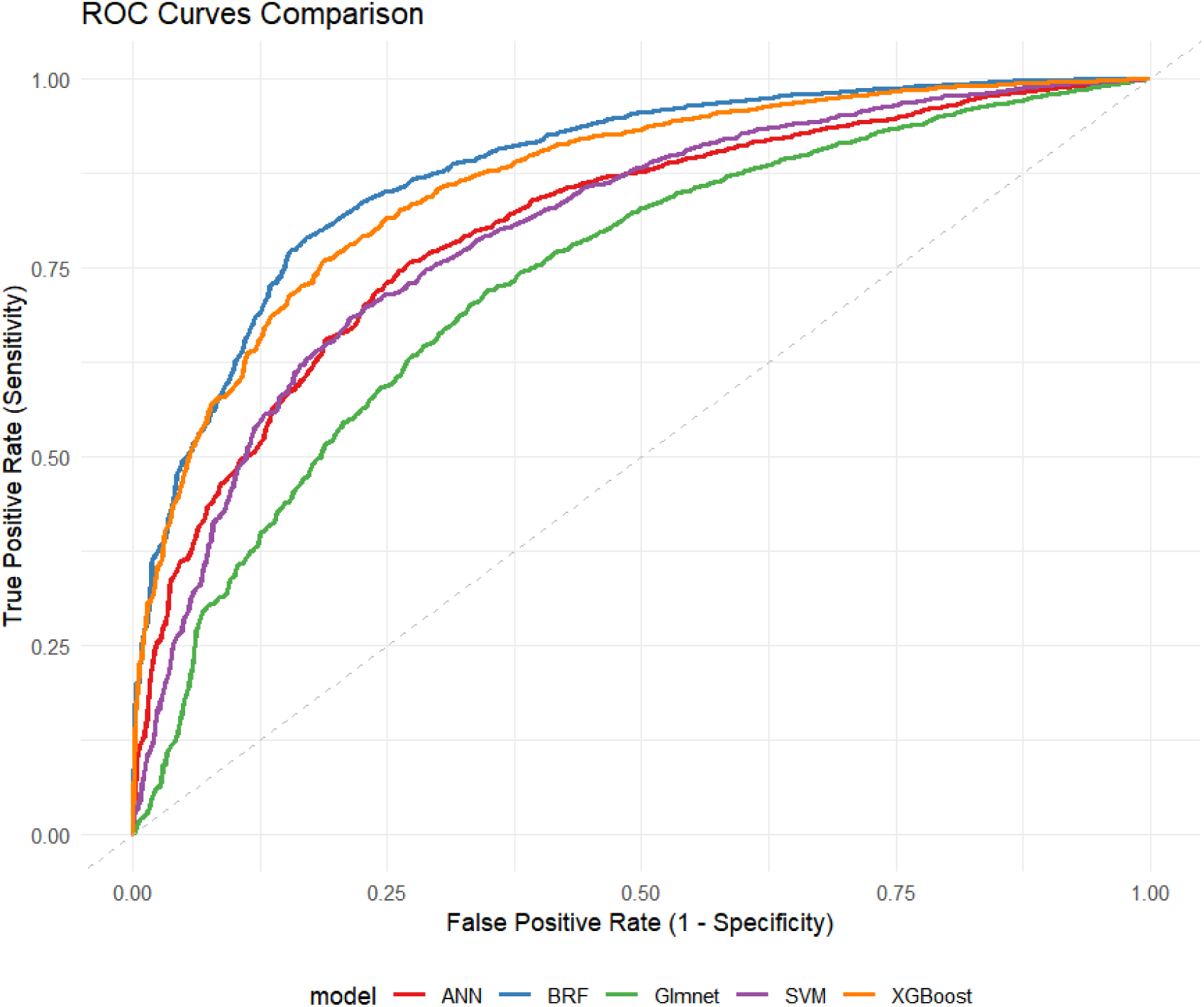
Receiver operating characteristic curves for the five machine learning models in predicting CKM syndrome.

**Figure 3B.**
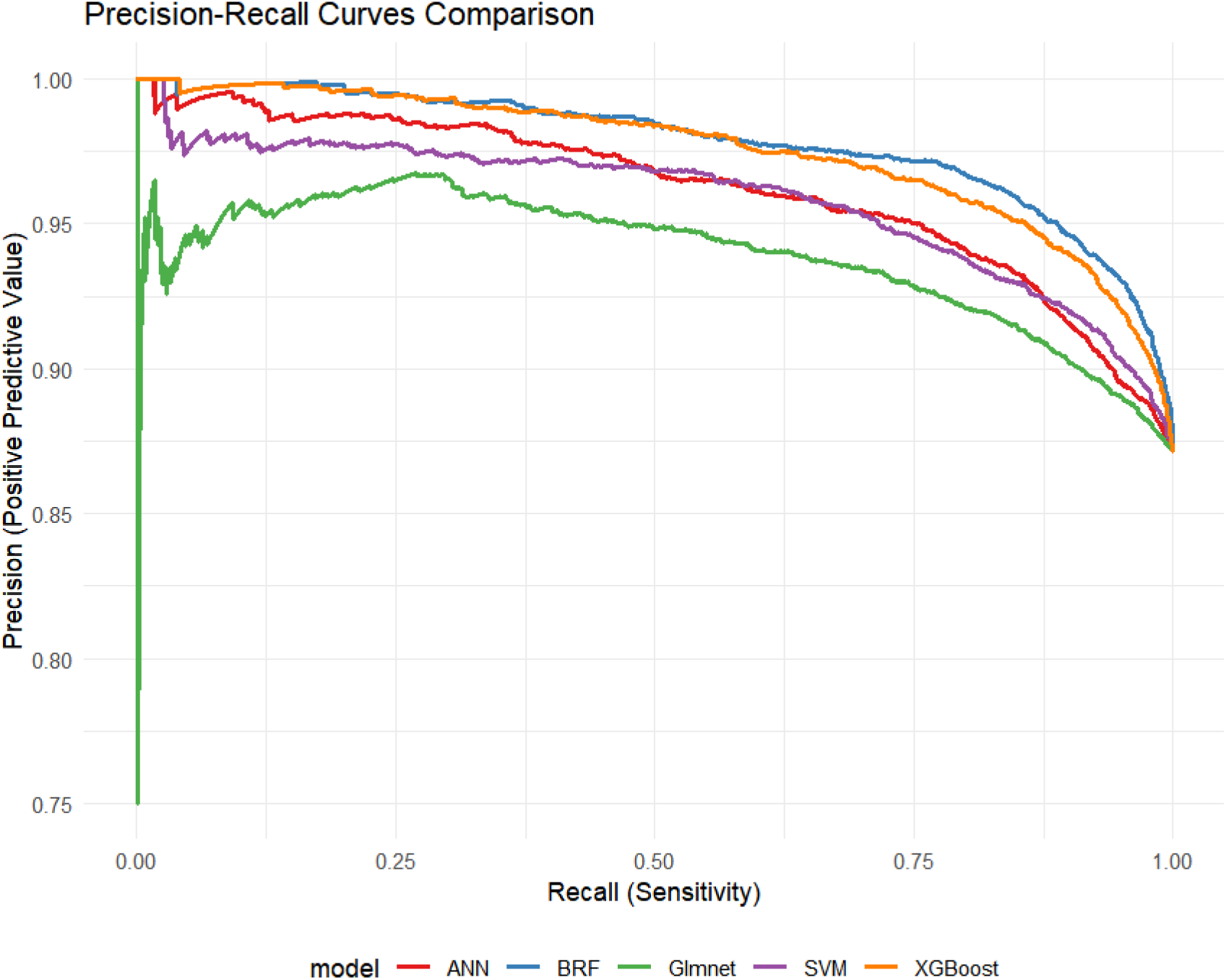
Precision-recall curves for the five machine learning models in predicting CKM syndrome.

**Table 4.** The characters of machine learning models in predicting CKM syndrome.

| Machine learning | Enhanced score | Accuracy | F-beta | Area under the ROC curve | Sensitivity | Specificity | Area under the PR curve |
| --- | --- | --- | --- | --- | --- | --- | --- |
| SVM | 0.770 | 0.773 | 0.851 | 0.831 | 0.929 | 0.523 | 0.913 |
| BRF | 0.862 | 0.851 | 0.898 | 0.912 | 0.940 | 0.651 | 0.954 |
| XGBoost | 0.893 | 0.848 | 0.885 | 0.922 | 0.846 | 0.852 | 0.960 |
| Glmnet | 0.711 | 0.729 | 0.819 | 0.748 | 0.884 | 0.680 | 0.855 |
| ANN | 0.783 | 0.761 | 0.829 | 0.813 | 0.835 | 0.695 | 0.906 |
| *p*-value | <0.001 | <0.001 | <0.001 | <0.001 | <0.001 | <0.001 | <0.001 |

SVM, support vector machine; BRF, balanced random forest; XGBoost,eXtreme gradient boosting; Glmnet, generalized linear models network; ANN, artificial neural network.

### 3.4 SHAP Value of Feature Importance in CKM Syndrome Prediction

The SHAP swarm plots highlighted the importance of various features in predicting CKM syndrome risk **(Figure 4A)**. Age emerged as the most significant predictor, strongly associated with increased CKM syndrome likelihood (SHAP value: 0.5351). Protective effects were identified for selenium (Se) (−0.1838), luteolin (−0.2019), total flavonoids (−0.1345), vitamin E (−0.1511), Mg (−0.1849), vitamin A (−0.1987), myricetin (−0.1483), vitamin C (−0.1862), kaempferol (−0.1546), zinc (Zn) (−0.1615), subtotal catechins (−0.1172), and total flavan-3-ols (−0.2073) **(Figure 4B)**. The force plot was generated to illustrate how these features influence model predictions **(Figure 4C)**. It illustrated the cumulative contribution of features toward a risk prediction, where protective dietary factors effectively counterbalance age risks.

**Figure 4A.**
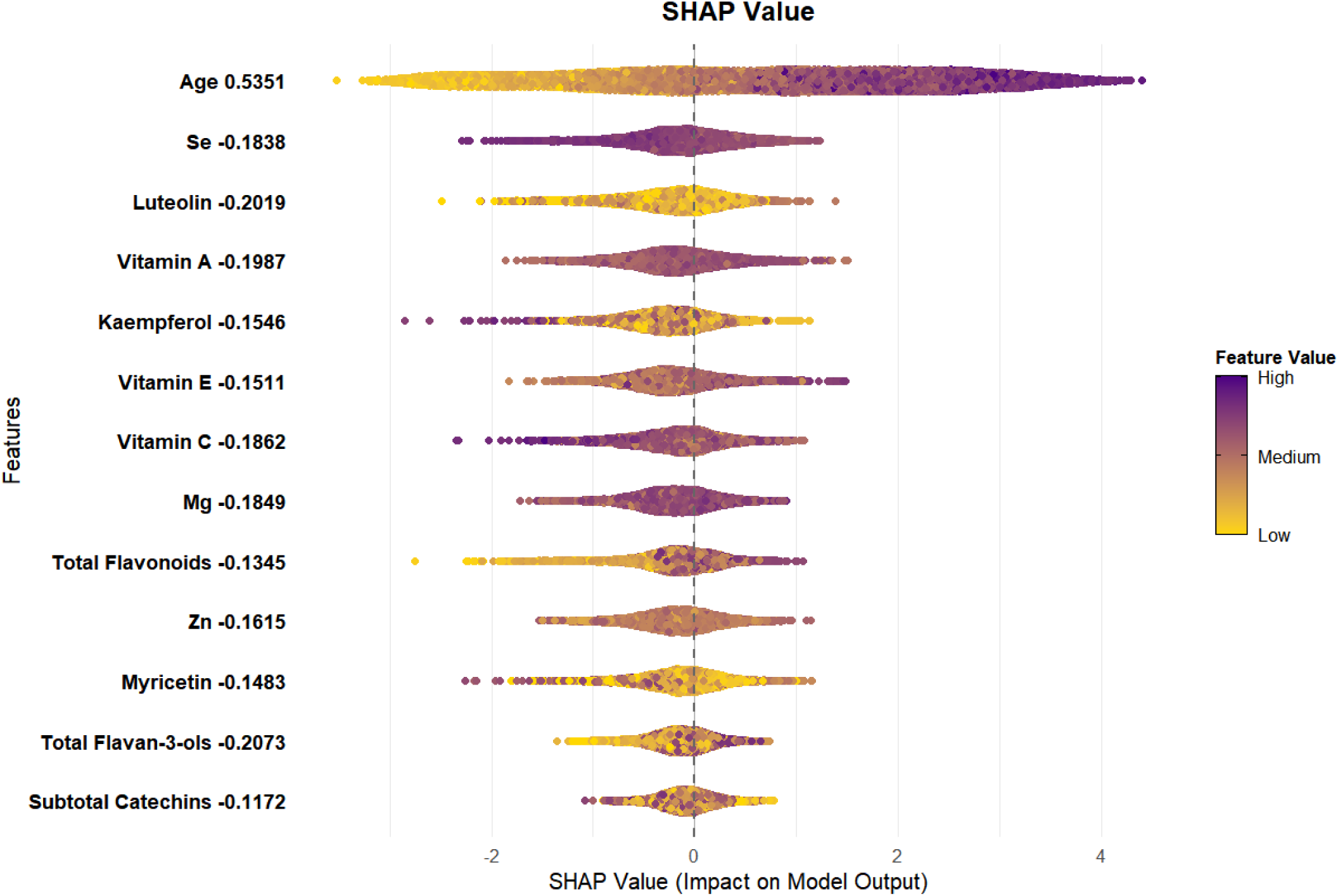
Swarm plot displaying SHAP value distributions for features within XGBoost model in CKM syndrome prediction.

**Figure 4B.**
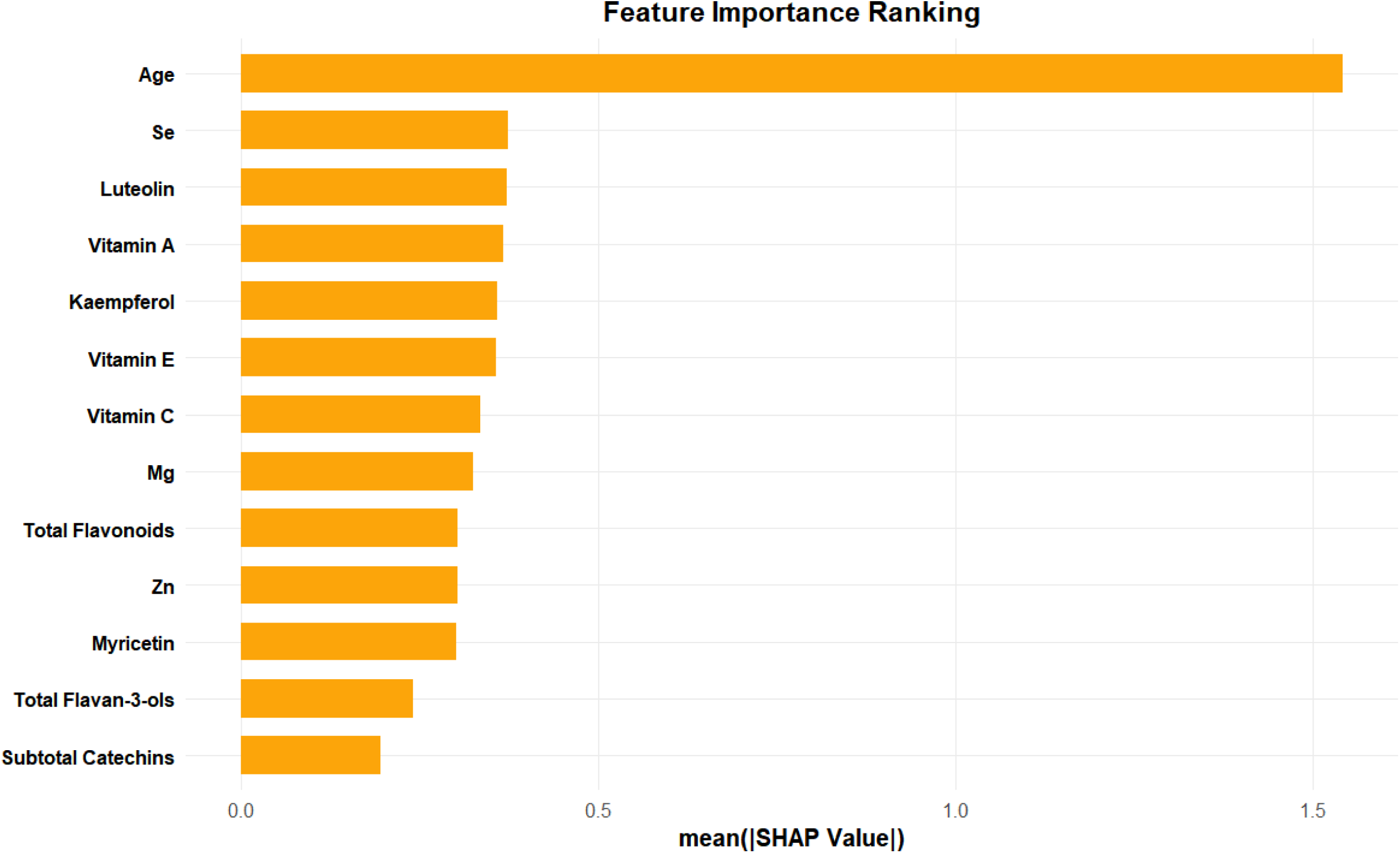
Waterfall plot of the features contributions to CKM syndrome prediction within XGBoost model.

**Figure 4C.**
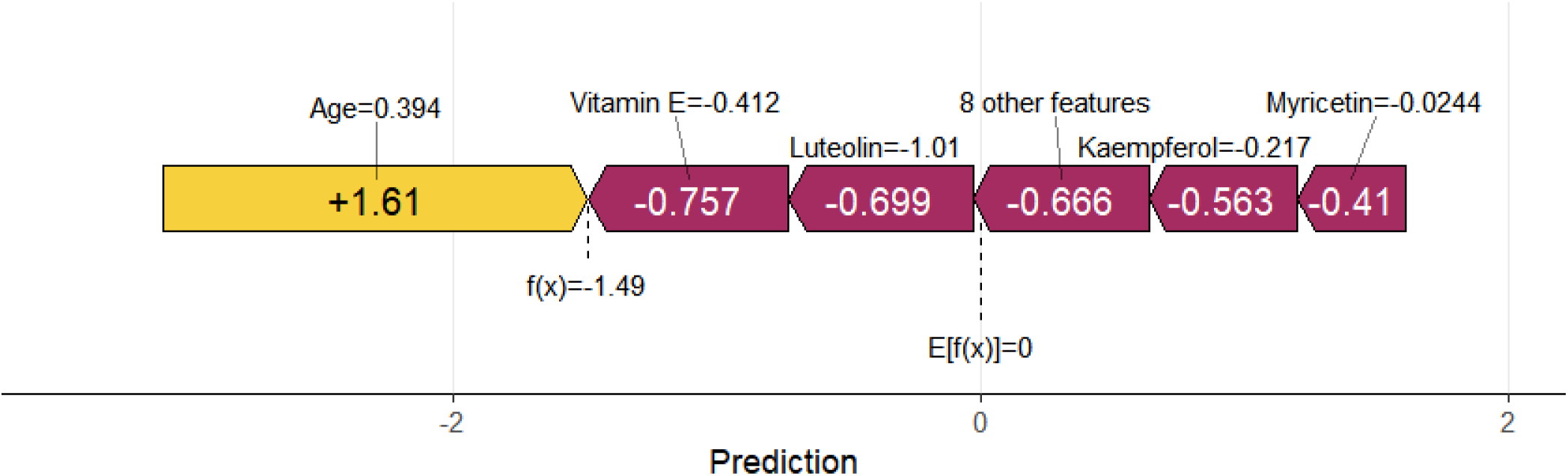
Force plot of the features contributions to CKM syndrome prediction within XGBoost model.

## 4. Discussion

In this study, we investigated the association between dietary antioxidant features and CKM syndrome using a hybrid ML approach with data from the NHANES database for the cycles of 2007-2008, 2009-2010, and 2017-2018 in the United States. Significant differences in socioeconomic and lifestyle factors were observed across various stages of CKM syndrome, including gender, race, education level, family income-to-poverty ratio, levels of moderate to vigorous physical activity, and current smoking status. XGBoost was identified as the optimal algorithm due to its superior predictive performance among the five assessed ML models. SHAP interpretability analysis indicated that age was the most significant risk factor, while selenium (Se), total flavonoids, vitamin E, Mg, vitamin A, myricetin, vitamin C, kaempferol, Zn, subtotal catechins, and total flavan-3-ols were key protective factors.

The application of ML in nutritional epidemiology has grown due to its ability to reveal complex, non-linear relationships between dietary features and disease. Research in related fields has highlighted the unique advantages of ML algorithms. SVM employed a kernel-based architecture to excel at high-dimensional classification tasks by identifying optimal hyperplanes[24]. This approach demonstrates strong performance with non-linear feature boundaries, making it particularly effective for analyzing antioxidant interactions within our study. BRF is an extension of traditional RF by using a balanced bootstrap sampling method to tackle class imbalance[25], which is essential for analyzing datasets with underrepresented groups, such as non-CKM syndrome individuals in our cohort. BRF effectively selects features despite skewed population distributions (CKM syndrome population: 90.8%) by overcoming the limitations of standard random forests. XGBoost demonstrated superior performance in handling collinear features in our dietary antioxidant datasets by minimizing feature-specific overfitting through its enhanced gradient-boosted framework[26], which includes parallelized tree construction and L1/L2 regularization. The penalized regression approach employed in Glmnet enhances interpretable variable selection by effectively distinguishing key dietary antioxidants from confounding noise. ANN was capable of capturing intricate interaction effects between dietary antioxidants and CKM syndrome. These models tackle complementary challenges in dietary antioxidant features-CKM syndrome modeling from dimensionality reduction to non-linear pattern recognition.

Our studies revealed that XGBoost was the most effective predictor of CKM syndrome among the five evaluated ML algorithms. Its gradient-boosting mechanism effectively managed collinear dietary antioxidants through regularization and minimized the overfitting risks associated with ANN. The computational efficiency of XGBoost surpassed that of AAN in our study, further supporting its practical utility in large-scale nutritional epidemiology. Moreover, the tree-based architecture of XGBoost inherently ranked feature importance, which corresponded well with the contributions of antioxidants as derived from SHAP analysis. This correspondence validated the biological relevance of XGBoost’s predictions and emphasized the additional value provided by SHAP interpretability. Although the sensitivity of the BRF was greater than that of XGBoost (0.940 vs. 0.893), its lower specificity (0.801 vs. 0.852) rendered it less suitable for clinical risk stratification, where minimizing false positives is paramount. XGBoost exhibited superior performance compared to SVM in capturing the non-linear relationships between antioxidants and CKM syndrome, resulting in enhanced predictive accuracy. Furthermore, SHAP visualizations for XGBoost revealed non-linear risk gradients associated with key antioxidants such as luteolin (SHAP value: –0.202), which aligned with the logistic regression model of linear trends (OR: 0.930). The integration of non-linear ML algorithms and linear logistic regression enhanced causal inference and exemplified the reliability of our findings. The application of ML models provided a robust methodological framework for identifying CKM syndrome predictors. The high AUC value of the XGBoost model indicated its effectiveness in classifying CKM syndrome risk, which is essential for the development of clinical tools aimed at early detection and intervention. Moreover, SHAP analysis further clarified the relative importance of features, highlighting the necessity for a multi-faceted approach to CKM syndrome management.

It marked a significant advancement over previous methods by systematically integrating five ML architectures with SHAP interpretability in this study, moving beyond traditional “black-box” approaches[27]. In contrast to previous studies that focused on single algorithms, our multi-model framework effectively reduced selection bias and identified robust dietary antioxidant features resilient to algorithmic variability. The process of cross-algorithm consensus harmonization significantly reduced the occurrence of false positives typically in univariate analyses. Additionally, we implemented SMOTE to address the significant group imbalance frequently observed in previous NHANES-based research. This methodological enhancement improved the generalizability of our model across all stages of CKM syndrome, although individuals in stage 4 constituted less than 5% of the cohort. Unlike conventional regression methods, the SHAP approach transcended limitations by elucidating the complex interactions through which antioxidants such as genistein mitigate age-related risks.

The findings from the SHAP value analysis indicated that age is the significant risk factor associated with CKM syndrome. Ageing is characterized by mitochondrial dysfunction, chronic low-grade inflammation, and endothelial impairment, all of which are critical for leading in leading to metabolic dysregulation and organ damage[28, 29]. Additionally, various socioeconomic and lifestyle factors also contributed significantly to CKM syndrome in our study. The higher prevalence of males within the CKM syndrome group **(Table 1)** reflected distinct sex-specific risk profiles. Males generally demonstrate a higher degree of visceral adiposity and androgen-induced insulin resistance, which heightens their vulnerability to cardiometabolic disorders. In contrast, premenopausal women benefit from the vasoprotective properties of estrogen, which aids in reducing oxidative stress and enhancing lipid metabolism. Gender plays a significant role in influencing the severity of CKM syndrome through intricate interactions involving sex hormones, the renin-angiotensin system, oxidative stress, inflammation, vascular disease, and insulin resistance[30]. Lower education levels and income-to-poverty ratios ≤3 **(Table 2)** highlighted the socioeconomic inequities associated with the burden of CKM syndrome progression. Individuals with limited education correlated with lower health literacy, which delayed preventive care and dietary modifications. Financial constraints further limited access to nutritious foods and healthcare services. Otherwise, the diversity of non-Hispanic black participants was prominent in the CKM syndrome stage group revealed systemic inequalities in resource allocation and chronic stress caused by discrimination. Reduced physical activity and smoking were linked to the progression of CKM syndrome, as supported by previous study[31]. Notably, the intake of daidzein and genistein exhibited a progressive decline of 73-76% from CKD stage 0 to stage 4. The results from ordinal logistic regression models further indicated that these two isoflavones were negatively associated with the progression of CKM syndrome. This finding underscores their dose-dependent protective relationship, emphasizing that the level of isoflavone intake is critical in influencing the progression of CKM syndrome.

The ordinal logistic regression analysis **(Table 3)** provided critical insights into the protective roles of dietary antioxidants at various stages of CKM syndrome progression, leveraging the ability to model linear trends across ordered outcomes (stages 0-4). Genistein, daidzein, delphinidin, apigenin, luteolin, and total flavones exhibited strong negative associations with CKD progression across all adjusted models. Notably, luteolin demonstrated the most significant protective effect, with a 7.0% reduction in odds for each mg/day increase in intake after adjusting for comprehensive demographic and lifestyle characteristics. Research has indicated that luteolin exerts its protective effects by suppressing NLRP3 inflammasome activation, regulating lipid metabolism through AMPK pathways, ameliorating pancreatic β-cell dysfunction via modulation of NF-κB expression, improving renal fibrosis by inhibiting TGF-β/Smad3 signaling pathways, and enhancing endothelial function through upregulation of nitric oxide synthase[32–36]. Similarly, kaempferol and total flavonols exhibited stage-dependent protective effects in Model 2 and Model 3, suggesting their broad-spectrum benefits across the entire CKM syndrome progression. The observed decrease in dietary antioxidant intake as CKD progresses may exacerbate oxidative stress—particularly among high-risk groups such as older males with limited health literacy. Notably, the negative association persisted even after controlling for socioeconomic factors, highlighting their biological significance beyond mere demographic differences.

The research methods employed in this study demonstrate several strengths that enhance the robustness and reliability of the findings. Firstly, a cross-sectional design was utilized, drawing on data from the NHAMES. The substantial sample size significantly increased the statistical power of the study, facilitating the identification of meaningful associations that may not be evident in smaller cohorts. Moreover, advanced statistical techniques were applied, including complex sequential ordinal logistic regression and machine learning algorithms, which provided a comprehensive understanding of the relationships between dietary antioxidant characteristics and stages of CKM syndrome. The incorporation of SHAP value analysis further improved model interpretability by elucidating the relative importance of various predictors. Collectively, these methodological strengths not only bolstered the validity of our results but also laid a foundation for future research aimed at exploring the intricate interactions between diet and the progression of chronic diseases.

The limitations of this study necessitate careful consideration. Firstly, the cross-sectional design inherently restricted the ability to establish causal relationships between dietary antioxidant features and CKM syndrome progression. Prospective cohorts with repeated dietary assessments are necessary to validate temporal relationships. Additionally, the self-reported dietary intake might introduce recall bias, potentially affecting the accuracy of the collected data. While the sample size of 5,349 participants was substantial, it might not adequately represent the broader population, particularly in terms of demographic diversity and geographical distribution. The lack of biochemical validation for observed associations could weaken the robustness of the findings. Lastly, variability among studies due to differences in data collection methods and participant characteristics may further limit the generalizability of these results. Addressing these issues in future research will be crucial for enhancing both the validity and clinical applicability of our findings.

## 5. Conclusion

In summary, this study elucidated the characteristics and risk factors associated with CKM syndrome progression, as well as the significant role of dietary antioxidants. The findings underscored the necessity of early screening and lifestyle interventions, positioning dietary antioxidant intake as a promising strategy to improve patient outcomes. Future longitudinal studies and clinical trials are warranted to validate these results and reinforce their implications for public health.

## Acknowledgments

We express our gratitude to all patients for their invaluable participation in this study.

## Statement of Ethics

This study received support from the Ethics Review Board of the U.S. National Center for Health Statistics. Informed written consent was obtained from all participants involved in the NAHNES survey.

## Author Contributions

X.D. and X.Y. and J.L. conceived of the presented idea and developed the study. X.D., D.R., F.Z., and G.X. assisted the data collection. X.D., F.Z., T.L., and Z.Z performed the analysis, X.D., G.X., and L.M. drafted the manuscript, J.L., D.R., Z.J and X.Y. assembled the figures. X.Y., D.R., N.Z., and Y.Z. revised the manuscript. All authors discussed the results and commented on the manuscript.

## Data Availability Statement

The datasets analyzed in this study are available in the article or supplementary material.

## Figure legends

**Table Supplement 1.**
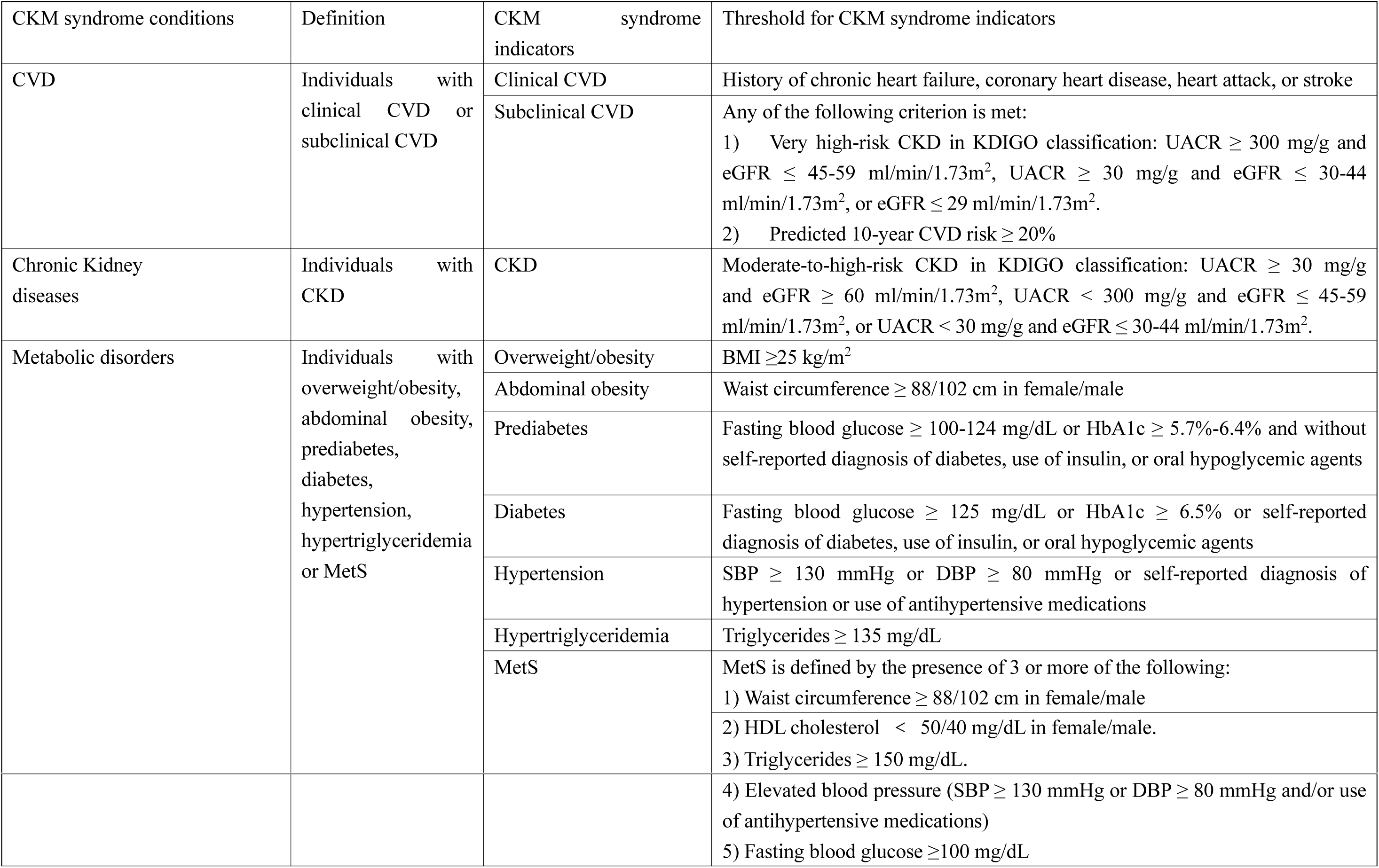
Definitions of CKM syndrome conditions.

**Table Supplement 2.**
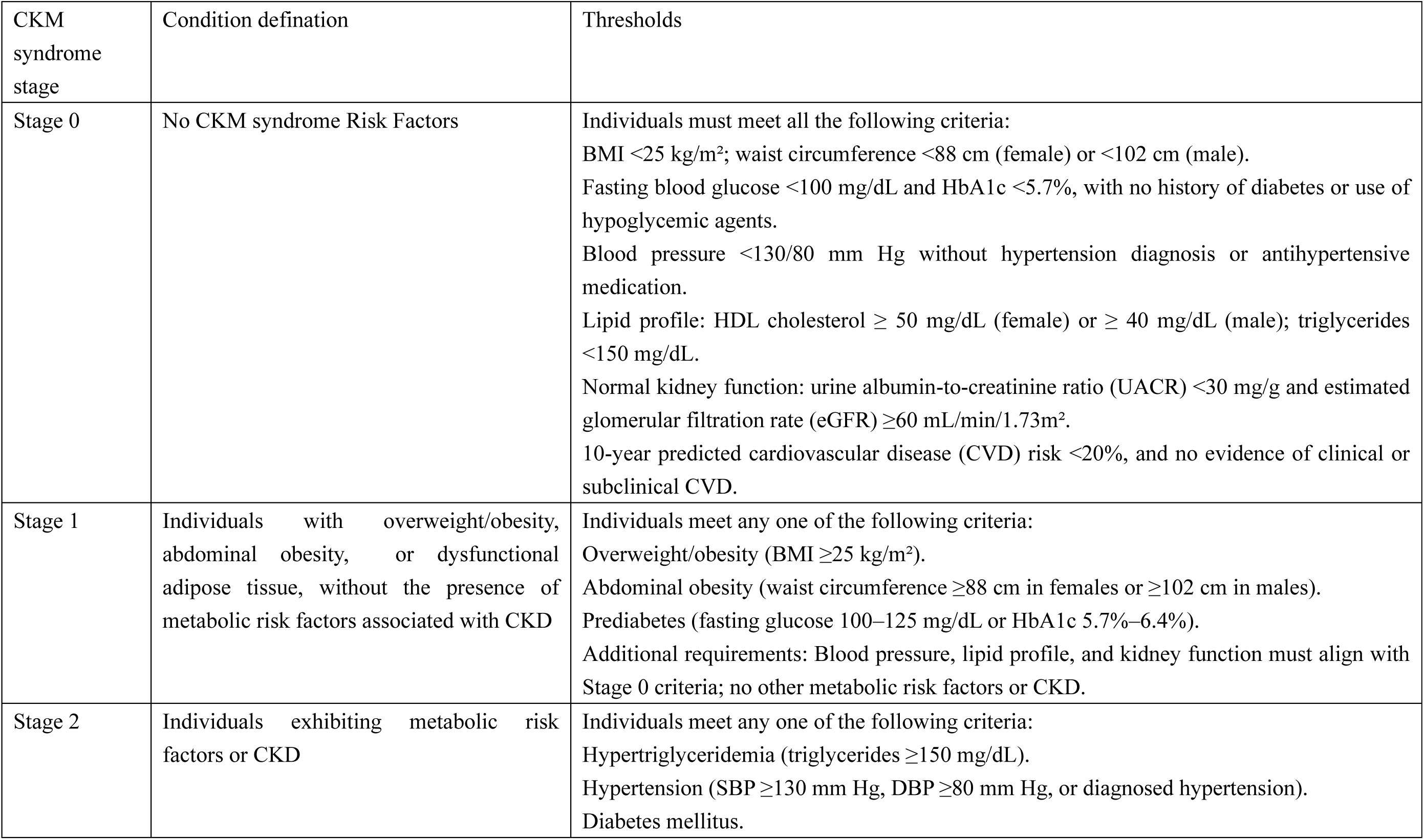
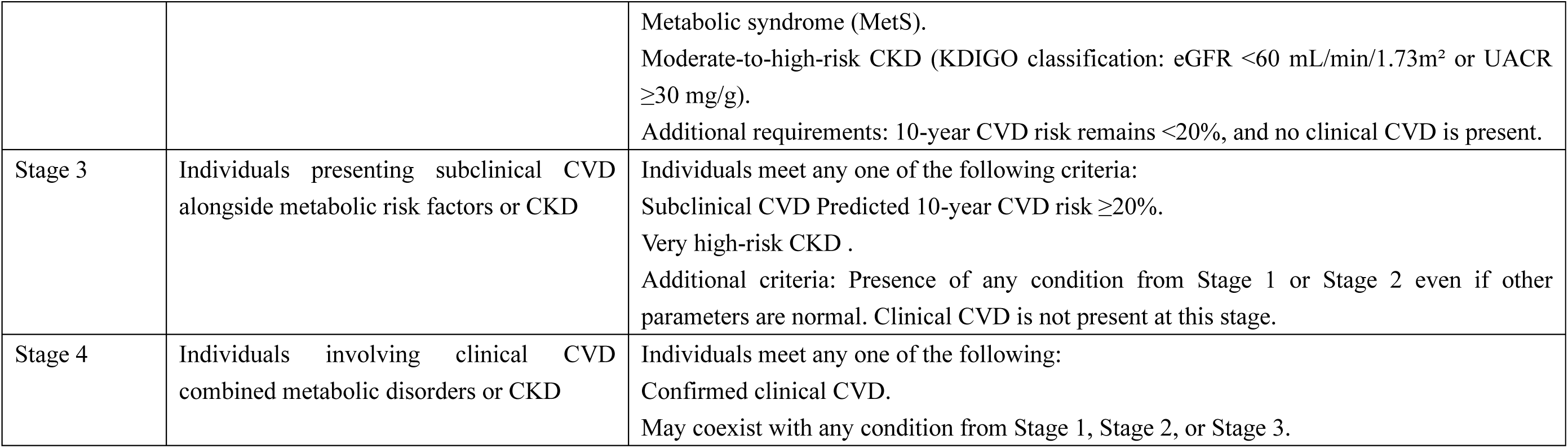
Definitions of CKM syndrome stages.

## References

1. Powell-Wiley TM, Poirier P, Burke LE, Després JP, Gordon-Larsen P, Lavie CJ, Lear SA, Ndumele CE, Neeland IJ, Sanders P et al: Obesity and Cardiovascular Disease: A Scientific Statement From the American Heart Association. Circulation 2021, 143(21):e984–e1010.

2. Rawshani A, Rawshani A, Franzén S, Sattar N, Eliasson B, Svensson AM, Zethelius B, Miftaraj M, McGuire DK, Rosengren A et al: Risk Factors, Mortality, and Cardiovascular Outcomes in Patients with Type 2 Diabetes. N Engl J Med 2018, 379(7):633–644.

3. Wright AK, Suarez-Ortegon MF, Read SH, Kontopantelis E, Buchan I, Emsley R, Sattar N, Ashcroft DM, Wild SH, Rutter MK: Risk Factor Control and Cardiovascular Event Risk in People With Type 2 Diabetes in Primary and Secondary Prevention Settings. Circulation 2020, 142(20):1925–1936.

4. Rangaswami J, Bhalla V, Blair JEA, Chang TI, Costa S, Lentine KL, Lerma EV, Mezue K, Molitch M, Mullens W et al: Cardiorenal Syndrome: Classification, Pathophysiology, Diagnosis, and Treatment Strategies: A Scientific Statement From the American Heart Association. Circulation 2019, 139(16):e840–e878.

5. Ndumele CE, Rangaswami J, Chow SL, Neeland IJ, Tuttle KR, Khan SS, Coresh J, Mathew RO, Baker-Smith CM, Carnethon MR et al: Cardiovascular-Kidney-Metabolic Health: A Presidential Advisory From the American Heart Association. Circulation 2023, 148(20):1606–1635.

6. Ostrominski JW, Arnold SV, Butler J, Fonarow GC, Hirsch JS, Palli SR, Donato BMK, Parrinello CM, O’Connell T, Collins EB et al: Prevalence and Overlap of Cardiac, Renal, and Metabolic Conditions in US Adults, 1999-2020. JAMA Cardiol 2023, 8(11):1050–1060.

7. Aggarwal R, Ostrominski JW, Vaduganathan M: Prevalence of Cardiovascular-Kidney-Metabolic Syndrome Stages in US Adults, 2011-2020. Jama 2024, 331(21):1858–1860.

8. Siri-Tarino PW, Sun Q, Hu FB, Krauss RM: Saturated fatty acids and risk of coronary heart disease: modulation by replacement nutrients. Curr Atheroscler Rep 2010, 12(6):384–390.

9. Niki E: Assessment of antioxidant capacity of natural products. Curr Pharm Biotechnol 2010, 11(8):801–809.

10. Schürks M, Glynn RJ, Rist PM, Tzourio C, Kurth T: Effects of vitamin E on stroke subtypes: meta-analysis of randomised controlled trials. Bmj 2010, 341:c5702.

11. da Silva A, Caldas APS, Pinto SL, Hermsdorff HHM, Marcadenti A, Bersch-Ferreira ÂC, Torreglosa CR, Weber B, Bressan J: Dietary total antioxidant capacity is inversely associated with cardiovascular events and cardiometabolic risk factors: A cross-sectional study. Nutrition 2021, 89:111140.

12. Mirmiran P, Hosseini-Esfahani F, Esfandiar Z, Hosseinpour-Niazi S, Azizi F: Associations between dietary antioxidant intakes and cardiovascular disease. Sci Rep 2022, 12(1):1504.

13. Mansour H, Slika H, Nasser SA, Pintus G, Khachab M, Sahebkar A, Eid AH: Flavonoids, gut microbiota and cardiovascular disease: Dynamics and interplay. Pharmacol Res 2024, 209:107452.

14. Liu P, Peng W, Hu F, Li G: Association between dietary intake of flavonoid and chronic kidney disease in US adults: Evidence from NHANES 2007-2008, 2009-2010, and 2017-2018. PLoS One 2024, 19(8):e0309026.

15. Guo XF, Ruan Y, Li ZH, Li D: Flavonoid subclasses and type 2 diabetes mellitus risk: a meta-analysis of prospective cohort studies. Crit Rev Food Sci Nutr 2019, 59(17):2850–2862.

16. Khan SS, Coresh J, Pencina MJ, Ndumele CE, Rangaswami J, Chow SL, Palaniappan LP, Sperling LS, Virani SS, Ho JE et al: Novel Prediction Equations for Absolute Risk Assessment of Total Cardiovascular Disease Incorporating Cardiovascular-Kidney-Metabolic Health: A Scientific Statement From the American Heart Association. Circulation 2023, 148(24):1982–2004.

17. Li J, Lei L, Wang W, Ding W, Yu Y, Pu B, Peng Y, Li Y, Zhang L, Guo Y: Social Risk Profile and Cardiovascular-Kidney-Metabolic Syndrome in US Adults. J Am Heart Assoc 2024, 13(16):e034996.

18. Stevens PE, Levin A: Evaluation and management of chronic kidney disease: synopsis of the kidney disease: improving global outcomes 2012 clinical practice guideline. Ann Intern Med 2013, 158(11):825–830.

19. KDIGO 2020 Clinical Practice Guideline for Diabetes Management in Chronic Kidney Disease. Kidney Int 2020, 98(4s):S1–s115.

20. Kwekha-Rashid AS, Abduljabbar HN, Alhayani B: Coronavirus disease (COVID-19) cases analysis using machine-learning applications. Appl Nanosci 2023, 13(3):2013–2025.

21. Fang W, Wu J, Cheng M, Zhu X, Du M, Chen C, Liao W, Zhi K, Pan W: Diagnosis of invasive fungal infections: challenges and recent developments. J Biomed Sci 2023, 30(1):42.

22. Erickson BJ, Korfiatis P, Akkus Z, Kline TL: Machine Learning for Medical Imaging. Radiographics 2017, 37(2):505–515.

23. Cao B, Zhang KC, Wei B, Chen L: Status quo and future prospects of artificial neural network from the perspective of gastroenterologists. World J Gastroenterol 2021, 27(21):2681–2709.

24. Bennett-Lenane H, Griffin BT, O’Shea JP: Machine learning methods for prediction of food effects on bioavailability: A comparison of support vector machines and artificial neural networks. Eur J Pharm Sci 2022, 168:106018.

25. Quist J, Taylor L, Staaf J, Grigoriadis A: Random Forest Modelling of High-Dimensional Mixed-Type Data for Breast Cancer Classification. Cancers (Basel) 2021, 13(5).

26. Dang X, Yang R, Jing Q, Niu Y, Li H, Zhang J, Liu Y: Association between high or low-quality carbohydrate with depressive symptoms and socioeconomic-dietary factors model based on XGboost algorithm: From NHANES 2007-2018. J Affect Disord 2024, 351:507–517.

27. Choi RY, Coyner AS, Kalpathy-Cramer J, Chiang MF, Campbell JP: Introduction to Machine Learning, Neural Networks, and Deep Learning. Transl Vis Sci Technol 2020, 9(2):14.

28. Amorim JA, Coppotelli G, Rolo AP, Palmeira CM, Ross JM, Sinclair DA: Mitochondrial and metabolic dysfunction in ageing and age-related diseases. Nat Rev Endocrinol 2022, 18(4):243–258.

29. Guo J, Huang X, Dou L, Yan M, Shen T, Tang W, Li J: Aging and aging-related diseases: from molecular mechanisms to interventions and treatments. Signal Transduct Target Ther 2022, 7(1):391.

30. Guldan M, Unlu S, Abdel-Rahman SM, Ozbek L, Gaipov A, Covic A, Soler MJ, Covic A, Kanbay M: Understanding the Role of Sex Hormones in Cardiovascular Kidney Metabolic Syndrome: Toward Personalized Therapeutic Approaches. J Clin Med 2024, 13(15).

31. Shang B, Yao Y, Yin H, Xie Y, Yang S, You X, Liu H, Wang M, Ma J: In utero, childhood, and adolescence tobacco smoke exposure, physical activity, and chronic kidney disease incidence in adulthood: evidence from a large prospective cohort study. BMC Med 2024, 22(1):528.

32. Lee MN, Lee Y, Wu D, Pae M: Luteolin inhibits NLRP3 inflammasome activation via blocking ASC oligomerization. J Nutr Biochem 2021, 92:108614.

33. Zhang L, Han YJ, Zhang X, Wang X, Bao B, Qu W, Liu J: Luteolin reduces obesity-associated insulin resistance in mice by activating AMPKα1 signalling in adipose tissue macrophages. Diabetologia 2016, 59(10):2219–2228.

34. Ge X, Liu T, Wang Y, Wen H, Huang Z, Chen L, Xu J, Zhou H, Wu Q, Zhao C et al: Porous starch microspheres loaded with luteolin exhibit hypoglycemic activities and alter gut microbial communities in type 2 diabetes mellitus mice. Food Funct 2025, 16(1):54–70.

35. Zhu Y, Liu R, Shen Z, Cai G: Combination of luteolin and lycopene effectively protect against the “two-hit” in NAFLD through Sirt1/AMPK signal pathway. Life Sci 2020, 256:117990.

36. Yang JT, Qian LB, Zhang FJ, Wang J, Ai H, Tang LH, Wang HP: Cardioprotective effects of luteolin on ischemia/reperfusion injury in diabetic rats are modulated by eNOS and the mitochondrial permeability transition pathway. J Cardiovasc Pharmacol 2015, 65(4):349–356.

